# Older adults’ beliefs about coping strategies for anxiety: A UK-based multicultural qualitative study informed by Leventhal’s Common-Sense Model of Self-Regulation

**DOI:** 10.64898/2026.03.28.26349595

**Authors:** Rasha Alkholy, Penny Bee, Rebecca Pedley, Karina Lovell

## Abstract

1

**Aim:** Older adults experiencing anxiety disorders, particularly those from minority ethnic backgrounds, are less likely to use formal mental health services compared to their younger counterparts. This UK multicultural qualitative study aimed to explore and compare beliefs underpinning coping strategies for anxiety among self-reporting White British, South Asian, African and Caribbean older adults, using Leventhal’s Common-Sense Model of Self-Regulation.

**Methods:** Individual semi-structured interviews were undertaken with 52 older adults aged 65 and over who self-reported (current or past) anxiety. Professional interpreters supported interviews with non-English-speaking participants (n=10). Eight public contributors collaborated on different aspects of the study. The Framework Method was used to manage and analyse the data.

**Findings:** The study drew on the perspectives of 27 older adults with distressing anxiety and 25 with non-distressing anxiety. Across all cultural groups, participants adopted different strategies to manage anxiety, the most prominent of which were self-help strategies. Help-seeking behaviour was influenced by a complex interplay of factors not recognised by Leventhal’s Common-Sense Model. Notably, older adults’ salient identities, rather than their cultural backgrounds, influenced their selection of coping strategies.

**Conclusions:** Interventions that empower older adults to use self-help strategies more effectively can serve as acceptable adjuncts to formal therapy. Nevertheless, addressing barriers to formal help-seeking is essential, particularly among those with a perceived need to seek help. No one model can depict the complexity of coping behaviours. While applying Leventhal’s Common-Sense Model yielded novel insights, it could not fully capture the motivational factors underlying participation in specific coping behaviours. To provide nuanced and accurate insights, cross-cultural research should acknowledge heterogeneity within groups rather than impose boundaries of purportedly homogeneous entities.

## 2 INTRODUCTION

Despite their negative impact on health [1] and healthcare costs [2], anxiety disorders remain largely underdiagnosed and undertreated. Older adults with anxiety disorders are less likely to use formal mental health services compared to their younger counterparts [3] - a trend that is particularly evident among those from minority ethnic backgrounds [4]. Furthermore, when older adults do seek help, they are less likely to receive treatment [5], and if they do, are more often offered psychotropic medications than psychological therapy [6].

Such disparities in mental healthcare use and treatment provision require extensive exploration if we are to achieve health equity. Unfortunately, research has predominantly overlooked factors underlying these disparities beyond sociodemographic and need factors [4]. One set of factors implicated in differential health services use in physical health problems are psychosocial factors, including people’s beliefs about illness and its treatment. These beliefs are central to Leventhal’s Common-Sense Model of Self-Regulation [7]. This model outlines how individuals conceptualise health threats, select coping actions and respond to feedback from their actions. It divides the behavioural process into three stages: a perceptual stage, in which a common-sense representation of the threat is developed (i.e., illness representations); a response stage, in which coping strategies are activated; an appraisal stage, in which coping strategies are evaluated for their success in achieving desired goals [8]. Coping strategies encompass self-help, help-seeking, taking medication or engaging in treatment and the action plans needed for individuals to implement them [9]. Similar to illness representations, beliefs about treatment (i.e., treatment representations) include the treatment’s identity (i.e., label and experiences when used), cause (i.e., mode of action), timeline (i.e., treatment duration, time to effect), consequences or adverse effects, perceived effectiveness to control or cure the illness [10] and emotional reactions [11]. A key aspect of the model is the need for coherence between an individual’s illness representation, their chosen coping strategies and their perception that their desired goals are being met [12].

In a previous analysis [13], we explored older adults’ beliefs and perceptions of anxiety (i.e., illness representations) and found that their understanding of anxiety lacked clarity - participants neither considered anxiety as a ‘normal’ part of ageing nor conceptualised it as an ‘illness’ trajectory. This ambiguity may help explain the low rates of mental health service utilisation in this population. However, to fully understand why older adults do not seek help for anxiety, despite experiencing distressing symptoms, it is necessary to look beyond illness representations alone and explore the beliefs that underpin their coping strategies. A cross-cultural examination is particularly important, as cultural norms and values can influence these beliefs [14]. Such insights may help us develop more acceptable services that are responsive to the needs of culturally diverse older adult populations. To our knowledge, no study has yet explored older adults’ beliefs about coping strategies for anxiety using Leventhal’s Common-Sense Model [10] (i.e., treatment representations).

## 3 METHODOLOGY AND METHODS

### 3.1 Study design

In this paper, we present a distinct analysis of primary data from a large, multicultural qualitative dataset [15] examining perceptions of anxiety and coping strategies among self-reporting White British, South Asian and African or Caribbean older adults in the UK. While a separate paper [13] reports an analysis focused specifically on older adults’ perceptions and representations of anxiety, the present paper addresses a different research question and offers a distinct conceptual contribution. Here, we explore participants’ beliefs about coping strategies through the lens of Leventhal’s Common-Sense Model [10]. Although both papers draw on the same dataset, they employ analytically independent thematic frameworks and address non-overlapping aspects of the data. We employed the generic qualitative approach described by Ritchie and Spencer (16); a pragmatic methodology that aligns with subtle realism and interpretivism.

### 3.2 Patient and Public Involvement and Engagement Group

Eight public contributors collaborated on different aspects of the study: five White British, two South Asian and one Caribbean, all living in the UK. Most of the public contributors were 65 years or over (n=7) and had lived experiences of common mental health problems (n=5). They coproduced the study materials, recruitment strategy and an arts-based public engagement workshop, served as Recruitment Champions, contributed to data interpretation and the final thematic framework, and co-authored a commentary on their involvement experience [13].

### 3.3 Ethical approval

The study received ethical approval from the University of Manchester Research Ethics Committee (Ref: 2021-10461-17766; Date: 05/02/2021).

### 3.4 Sampling and Recruitment

Participants were eligible if they were 65 years or over; self-identified as being “worriers”, had experienced or were experiencing anxiety or “stress”; self-identified as being White British, South Asian (Indian, Pakistani, Bangladeshi), African or Caribbean; and living in the UK. Individuals were excluded if they were receiving mental health crisis management. We used purposive sampling to ensure adequate representation across the three cultural groups and snowball sampling to maximise our reach to eligible participants.

Data were primarily collected during the national COVID-19 lockdown. We asked relevant gatekeepers (e.g., Older Black Africans Day Opportunities, the African Caribbean Care Group, Khush Amdid), research networks (e.g., Centre for Applied Dementia Studies at the University of Bradford) and our public contributors to circulate the study details to their contacts. Study flyers were posted on study-specific social media platforms (X/Twitter, Facebook), as well as on the websites and newsletters of the University of Manchester and third-sector and charity organisations (e.g., Anxiety UK, Parkinsons UK). Physical study flyers were distributed in venues like local churches, mosques and pharmacies to reach digitally excluded older adults.

Due to concerns about the acceptability of the term “anxiety” among older adults, our public contributors recommended the use of different terms in the study flyers, such as “stress” and “worrier” and their synonyms in Urdu, Hindi and Bengali.

Older adults interested in the study were encouraged to get in direct contact with the researcher using the contact details provided in the study advert. Older adults who knew about the study through gatekeepers or public contributors and explicitly stated that they would prefer to be contacted by the researcher or required interpretation services were asked for their consent to share one method of contact.

During initial contact, the researcher provided a brief verbal explanation of the study and screened potential participants for their eligibility to take part. Potential participants were told that this study aimed to understand people’s experiences of anxiety that persisted over long periods and impacted their daily functioning or caused significant distress. Those who self-reported anxiety, either currently or in the past, based on this broad description, and those who had a clinical diagnosis of anxiety were eligible to participate.

Eligible participants were provided with a ‘Research Package’ including the Participant Information Sheet, consent form and three questionnaires: a sociodemographic form to capture basic sociodemographic data, and the Geriatric Anxiety Scale [17] and Geriatric Depression Scale-Short Form [18] to screen for current symptoms of anxiety and depression, respectively.

The study advert, Participant Information Sheet, the Geriatric Anxiety Scale [17] and the consent form were translated into Urdu, Hindi and Bengali with all translations proofread and verified by certified independent translators. The Urdu study materials were also reviewed by an Urdu-speaking public contributor to ensure linguistic appropriateness and cultural acceptability. The English [18], Urdu [19], Hindi [20] and Bengali [21] versions of the Geriatric Depression Scale-Short Form are available from the respective source publications. The Geriatric Anxiety Scale total score was only used to evaluate the effectiveness of our inclusive recruitment strategy.

Upon receipt of the study materials, participants were invited to an introductory meeting with the researcher to review the Participant Information Sheet and address questions. Professional interpreters facilitated introductory meetings with non-English-speaking participants. This was particularly important for non-English-speaking participants who communicated in languages other than Urdu, Hindi or Bengali, as additional time was required to thoroughly review and interpret the information sheet.

Participants were informed that participation was voluntary and that, even after providing consent, they retained the right to withdraw. To mitigate potential coercion, participants identified and approached by public contributors or gatekeepers were informed that their decision to participate or not would remain confidential. All participants were afforded at least 24 hours to consider their consent for participation prior to scheduling their interviews.

### 3.5 Data collection

Participants took part in two interviews: a brief initial interview in which verbal consent and answers to the three questionnaires were audio-recorded, followed by a semi-structured interview ranging from 38 to 160 minutes.

In accordance with COVID-19 national lockdown restrictions that were in place during the data collection period, informed consent was obtained verbally rather than in written form. Verbal consent was audio-recorded and subsequently documented in a secure consent log to ensure an auditable record. This consent procedure, including the use of audio-recorded verbal consent, was reviewed and approved by the University of Manchester Research Ethics Committee and complied with institutional COVID-19 guidance for remote research at the time.

The semi-structured interviews followed a topic guide, coproduced with the public contributors, based on Leventhal’s Common-Sense Model [10]. The guide included five open-ended questions with follow-up probes to explore participants’ conceptualisation of anxiety, their self-help strategies, help-seeking behaviours and beliefs about treatments, perceived barriers and enablers to mental health services use, and the outcomes they used to appraise different strategies. We iteratively adapted the topic guide to include new topics or perspectives introduced by the participants. In this paper, we report only the findings related to coping strategies (i.e., self-help strategies, help-seeking behaviours, beliefs about treatments, barriers and enablers to service use, appraisal outcomes).

Ten interviews were conducted in non-English languages, including Wolof (n= 1), Luganda (n= 2), Urdu (n= 4), Punjabi (n= 2) and Sylheti (n= 1). Twelve interviews were conducted in person (seven at participants’ homes, three in community organisations, two in university facilities), 27 by telephone and 13 virtually (Zoom University account). Data collection took place in the UK over a 13-month period (23 April 2021 to 30 May 2022). All participants received a £30 gift voucher in recognition of their time.

### 3.6 Data analysis

We used the Framework Method [16] to manage and analyse the data. This comparative form of thematic analysis involves using an organising framework of deductively and inductively derived themes as a scaffold against which data are analysed. Its hallmark feature is its matrix output consisting of rows (cases), columns (codes) and cells of ‘summarised data’, providing a structure for systematically reducing and analysing the data both by case and code. The matrix output allows a bird’s-eye view of the entire dataset while maintaining the integrity of each participant’s account [22], a feature particularly relevant to our study as we wanted to compare participants’ perspectives both across and within the three cultural groups. Leventhal’s Common-Sense Model [7] served as the a priori organising framework that guided the initial deductive analysis. We also used inductive analysis to extend and refine the thematic framework, allowing the identification of new dimensions not recognised by the original model.

RA familiarised herself with the transcripts, identified initial codes and developed a preliminary thematic framework, which was subsequently refined and applied to the whole dataset. The research team (KL, PB, RP, RA) met regularly throughout this process to discuss and verify coding, refine the thematic framework and review matrix outputs. Our public contributors reviewed selected extracts of anonymised quotes and provided input on data interpretation and the final thematic framework. Any discrepancies were resolved to ensure consensus among the wider group. Data management was supported by NVivo 12 Plus.

We agreed that data saturation was reached when codes, themes, linkages and interpretations were judged to be sufficiently developed, conceptually dense and coherent to address the study’s objectives. Data saturation was reached after interviews with 14 White British, 16 South Asian and 15 African or Caribbean participants. To validate this assessment, we conducted one to two additional interviews with the African/Caribbean and White British groups, respectively. A further four interviews were conducted with the South Asian group to ensure adequate linguistic diversity within the sample.

The interviewer (RA) is a female physician with training in qualitative research and had no prior relationship with any participant. Any safety concerns that arose during data collection were discussed with the lead clinician (KL). RA used reflexive notes to document assumptions, decisions and dynamics that may have impacted the research process, highlighting personal, interpersonal, methodological and contextual issues. An audit trail was used to record all research steps.

## 4 FINDINGS

### 4.1 Participant characteristics

We interviewed 52 older adults: 16 White British, 20 South Asian and 16 African or Caribbean. The mean age of the participants was 73 years, and most of them were females.

Although all participants self-reported current or past experiences of anxiety, our sample comprised two distinct subgroups: those who, at the time of experiencing anxiety, experienced significant distress (defined as a subjective sense of emotional pain accompanied by an inability to cope) and/or functional impairment (defined as limitations in important areas of functioning attributable to anxiety) and those who did not. We refer to the first group as participants ‘with distressing anxiety’ and the latter as participants ‘with non-distressing anxiety’. This study draws on the perspectives of 27 older adults with distressing anxiety and 25 with non-distressing anxiety. The characteristics of the participants are outlined in **Table 1**.

**Table 1:** Participant characteristics.

| Participant characteristic | White British (n= 16) | South Asian (n= 20) | African/Caribbean (n= 16) | Total (n= 52) |
| --- | --- | --- | --- | --- |
| Age (years), mean (SD) | 73.6 (6.6) | 70.9 (6.6) | 75.4 (8.4) | 73.1 (7.3) |
| Age range (years) | 66 - 91 | 65 - 86 | 66 - 88 | 65 - 91 |
| **Sex** |  |  |  |  |
| Female, n (%) | 10 (62.5) | 14 (70.0) | 13 (81.3) | 37 (71.2) |
| Male, n (%) | 6 (37.5) | 6 (30.0) | 3 (18.7) | 15 (28.2) |
| Education (years), mean (SD) | 14.4 (3.7) | 12.9 (6.8) | 13.7 (4.5) | 13.6 (5.3) |
| **Marital status** |  |  |  |  |
| Single | 0 | 0 | 2 | 2 |
| Married | 10 | 11 | 4 | 25 |
| Separated | 0 | 0 | 3 | 3 |
| Divorced | 3 | 2 | 3 | 8 |
| Widowed | 2 | 7 | 4 | 13 |
| Prefer not to say | 1 | 0 | 0 | 1 |
| **Language used** |  |  |  |  |
| English | 16 | 13 | 13 | 42 |
| Non-English | 0 | 7 | 3 | 10 |
| • Urdu | 0 | 4 | 0 | 4 |
| • Punjabi | 0 | 2 | 0 | 2 |
| • Sylheti | 0 | 1 | 0 | 1 |
| • Luganda | 0 | 0 | 2 | 2 |
| • Wolof | 0 | 0 | 1 | 1 |
| **Living status** |  |  |  |  |
| Living alone | 4 | 4 | 9 | 17 |
| Living with others | 12 | 16 | 7 | 35 |
| **Needs help in performing daily activities** |  |  |  |  |
| Always | 2 | 3 | 0 | 5 |
| Most of the time | 1 | 3 | 1 | 5 |
| Sometimes | 2 | 2 | 1 | 5 |
| Rarely | 2 | 2 | 2 | 6 |
| Never | 9 | 10 | 12 | 31 |
| **Medical problems** |  |  |  |  |
| Yes | 16 | 18 | 13 | 47 |
| No | 0 | 2 | 3 | 5 |
| **Participants with distressing anxiety** | **9** | **10** | **8** | **27** |
| Currently experiencing symptoms suggestive of anxiety based on GAS total score <sup>A</sup> |  |  |  |  |
| • Yes | 8 | 9 | 6 | 23 |
| • No | 1 | 1 | 2 | 4 |
| Currently experiencing symptoms of depression based on GDS-SF score <sup>B</sup> |  |  |  |  |
| • No depression | 2 | 3 | 6 | 11 |
| • Suggestive of depression | 5 | 2 | 2 | 9 |
| • Almost always depression | 2 | 5 | 0 | 7 |
| Sought help from healthcare services for mental health concerns <sup>C</sup> |  |  |  |  |
| • Yes | 9 | 6 | 6 | 21 |
| • No | 0 | 4 | 2 | 6 |
| Has a clinically diagnosed mental health condition (i.e., formal diagnosis by a healthcare professional) |  |  |  |  |
| • None | 4 | 6 | 7 | 17 |
| • Anxiety | 1 | 1 | 0 | 2 |
| • Depression | 1 | 1 | 0 | 2 |
| • Anxiety and other comorbidities (depression, post-traumatic stress disorder) | 3 | 2 | 1 | 6 |
| Received treatment for anxiety (irrespective of formal diagnosis) |  |  |  |  |
| • Medication | 6 | 4 | 3 | 13 |
| • Psychotherapy | 5 | 1 | 4 | 10 |
| **Participants with non-distressing anxiety** | **7** | **10** | **8** | **25** |
| Currently experiencing symptoms suggestive of anxiety based on GAS total score <sup>A</sup> |  |  |  |  |
| • Yes | 6 | 3 | 2 | 11 |
| • No | 1 | 7 | 6 | 14 |
| Currently experiencing symptoms of depression based on total GDS-SF score <sup>B</sup> |  |  |  |  |
| • No depression | 7 | 10 | 8 | 25 |
| Sought help from healthcare services for mental health concerns <sup>C</sup> |  |  |  |  |
| • Yes | 2 | 3 | 2 | 7 |
| • No | 5 | 7 | 6 | 18 |
| Has a clinically diagnosed mental health condition (i.e., formal diagnosis by a healthcare professional) |  |  |  |  |
| • None | 7 | 8 | 7 | 22 |
| • Anxiety and other comorbidities (depression) | 0 | 0 | 1 | 1 |
| • Other mental health problems (bipolar disorder, obsessive compulsive disorder) | 0 | 2 | 0 | 2 |
| Received treatment for anxiety (irrespective of formal diagnosis) |  |  |  |  |
| • Medication | 0 | 0 | 0 | 0 |
| • Psychotherapy | 1 | 0 | 0 | 1 |

GAS= Geriatric Anxiety Scale; GDS-SF= Geriatric Depression Scale-Short Form; n=number; SD=standard deviation.
<sup>A</sup> Geriatric Anxiety Scale total score: > 9 currently experiencing symptoms suggestive of anxiety; ≤ 9 currently not experiencing symptoms suggestive of anxiety [23].
<sup>B</sup> Geriatric Depression Scale-Short Form total score: ≤5 no depression; 6-10 suggestive of depression; ≥ 11 almost always depression [18].
<sup>C</sup>= Any contact with formal healthcare services, including primary, secondary or tertiary services, and private or public services, for mental health reasons.

### 4.2 Thematic overview

We developed a thematic framework **(Fig 1)** incorporating a priori themes derived from Leventhal’s Common-Sense Model [10], against which data were deductively analysed, and potential new themes identified via inductive thematic analysis. These new themes include factors underlying the motivation to take action (or barriers and enablers to help-seeking) and beliefs about treatment providers. Beliefs about medications and psychotherapy were generally aligned with the a priori treatment-related dimensions proposed by Leventhal’s Common-Sense Model [10]. Additional new treatment-related beliefs were identified: conflicts with higher-order goals, compensatory behaviours and beliefs about factors which may determine the effectiveness of therapy.

**Fig 1:**
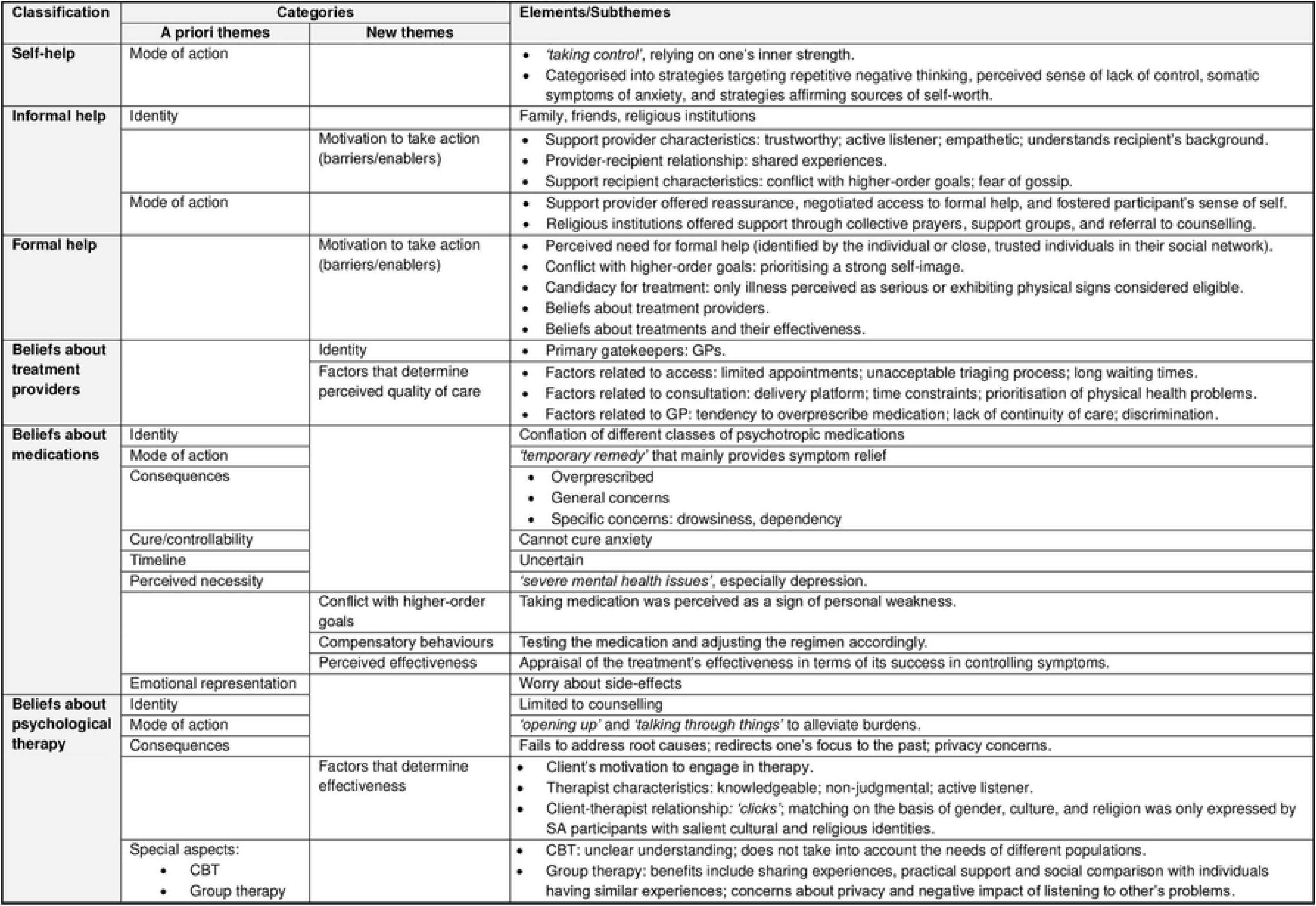
Thematic framework demonstrating older adults’ beliefs about coping strategies for anxiety

#### 4.2.1 Self-help

Across all cultural groups, participants emphasised the importance of *‘taking control’* and relying on their inner strength to manage their mental health concerns before seeking any form of support. *‘Self-help’* represented a toolkit of strategies used by individuals as a first-line, and sometimes the only, approach to coping with mental health concerns.

> *‘It’s kind of, you know, taking steps yourself…Anything that comes out of your own is much better than any intervention by an external person, you know.’ (South Asian male; non-distressing anxiety, used self-help only)*

Self-help strategies were categorised according to their primary target into strategies targeting repetitive negative thinking patterns like worry and rumination, perceived sense of lack of control, somatic symptoms of anxiety and strategies affirming sources of self-worth.

##### Strategies targeting repetitive negative thinking

Distraction was the most frequently cited self-help strategy across all cultural groups. The participants engaged in activities such as walking, watching TV and exercising, so that they were *‘not sitting in the house and dwelling on things.’* These activities helped them shift their focus from negative thoughts to less taxing ones. However, their effect was temporary.

##### Strategies targeting symptoms of anxiety

These were mainly behavioural strategies used by participants who attributed somatic symptoms, such as hyperventilation and gastrointestinal disturbance, to anxiety. They included breathing exercises and drinking soothing, warm fluids.

##### Strategies targeting a perceived sense of lack of control

Participants believed their anxiety was caused by unmodifiable stressors beyond their control, including progressive illness, cumulative social losses and traumatic or discriminatory experiences. This lack of control over one’s life was difficult to accept, especially among those with distressing anxiety.

> *‘Things you can’t logically, you can’t control. I can control what I do today, I can control what I do tomorrow. But you can’t change the past can you? But I couldn’t seem to accept this, and I’d dwell on it.’ (White British male; distressing anxiety, sought formal help, but refused treatment)*

Participants with salient religious identities, mainly from the African/Caribbean and South Asian groups, were able to resign their desire to be in control to a higher power who would *‘not forsake*’ them, but rather guide and protect them. Religious prayers offered a chance to connect and converse with God, thus providing comfort and reassurance.

> *‘But when you start a day and you pray and you commit everything to him [God], you go out and you have, it’s almost like you have an authority, you have a confidence, I’ve handed over everything to him and now he is in control and I am just going to be listening for God’s guidance.’ (African/Caribbean female; distressing anxiety, used self-help only)*

##### Strategies affirming sources of self-worth

Across all cultural groups, participants employed diverse strategies that shared a common underlying goal of maintaining an overarching positive view of the self. These strategies include self-affirming reflections, engagement in meaningful and purposeful activities, downward social comparison and involvement in social groups, particularly those one strongly identifies with. When participants perceived an unmodifiable stressor (e.g., progressive illness) as a threat to a specific aspect of their self-concept (i.e., independence), they leaned on and reinforced other unaffected aspects of their self (i.e., personal strengths and achievements, relationships and group memberships).

African/Caribbean participants tended to use self-affirming reflections as a reminder of their diverse and strong personal qualities instead of focusing on threatened aspects of their identity.

> *‘You have to tell yourself, “I am great, I am a wise woman. Yes, I am a grandmother, I am a professional, these are all roles, but they are not who I am. I am a wise woman.” Write it on the wall, say it to yourself, it is conditioning.’ (African/Caribbean female; distressing anxiety, sought formal help, but refused treatment)*

Participants from all cultural groups engaged in various activities, such as sewing, gardening and exercise, with the main goal of distracting themselves. However, only those participants who set achievable goals and focused on outcomes felt a sense of accomplishment that boosted their self-esteem when they achieved those outcomes.

> *‘…that [gardening] was really great for my mental health because I was nurturing these plants, oh so proud, and potatoes, I grew potatoes as well.’ (African/Caribbean female; distressing anxiety, sought formal help, received counselling and medication)*

Participants tended to seek comparisons with those who were worse off to feel better about themselves. However, for those with salient religious identities, downward social comparison seemed to be an inherent part of their belief.

> *‘Because when you listen to others it will just help you with your situation. If you are a Christian, a Muslim or somebody you believe in God, you will just go back to the comfort of your house and just give thanks to God.’ (African/Caribbean male; non-distressing anxiety, used self-help only)*

Participants also leaned on their relationships and group memberships to buffer the effect of threats to other aspects of their identity. Across all cultural groups, participants tended to join social groups based on age, activity (e.g., singing), disease (e.g., diabetes) or social roles (carers’ groups) to make friends, engage in activities and obtain practical advice. Only one participant was aware of social prescribing services, linking people to community organisations, but did not use them.

Although these groups offered social support, it was group memberships founded on shared, salient identities that fostered a sense of belonging, purpose and meaning. This pattern was evident among South Asian and African/Caribbean participants with salient religious and cultural identities, both of which were strongly *‘interlinked’.* African/Caribbean participants regarded in-group members who shared their religious beliefs and cultural background as an extension of their self. For these participants, the Church was not only a place of worship; it was a family.

> *‘I know the church and the pastor…you become one family, all the members are sisters and brothers, yeah.’ (African/Caribbean female; distressing anxiety, sought formal help, received counselling)*

South Asian women with salient religious or cultural identities explained how joining South Asian women-only social groups allowed them to connect with others who shared their cultural traditions, religious beliefs and language while respecting deeply rooted cultural norms and expectations.

> *‘Yes, that’s the reason that [name of social group] came, isn’t it? You bringing them [South Asian women] out from their understanding and, you know, mixing with people, that’s how important going out is how important, how helpful for your mental health is seeing when, because especially Asian community our community, women are left alone, isn’t it?’ (South Asian female; non-distressing anxiety, used self-help only)*

#### 4.2.2 Informal help (i.e., family, friends, religious institutions) for mental health concerns

Family and friends represented valuable social networks that participants turned to when experiencing mental health concerns. However, only a select few within these networks were trusted to provide support. These individuals understood the participant’s background and could relate to what they were going through because of their shared experiences, *‘we’re all in the same boat’.* They offered reassurance, encouragement and practical support when needed and, in some cases, negotiated the participant’s access to formal help. These people *‘give you strength’* - they fostered the participants’ sense of value.

> *‘They [grandchildren] understand very much, yes, and they even tell me, they told me, “We lost our grandpa, now we don’t want to lose you, so you have to be strong and conquer.” (African/Caribbean female; distressing anxiety, received informal help)*

Talking in itself had a cathartic effect.

> *‘If I didn’t tell anyone then it will damage, you know, I’m thinking, thinking, thinking so much and my heart, I’m not happy. When I tell someone like my sister or my friend then after that I’m okay, that’s okay.’ (South Asian female; distressing anxiety, sought formal help, received medication)*

Participants sought support from friends rather than close family members when they felt *‘it’s probably easier to talk to somebody outside’,* especially if they did not want to *‘burden’* their families or felt their families were *‘dismissive’*.

However, not all participants sought support from family or friends. Across all cultural groups, participants who valued maintaining a *‘strong’* persona were *‘embarrassed’* about seeking support. Similarly, those who prioritised projecting a *‘positive’* image avoided seeking help because they believed that people who were *‘always moaning about something’* were shunned by others.

> *‘No, no, I don’t talk to many people about my feelings because it’s always people ringing me up to ask me for advice, most of my friends and, you know…cause I’m like the matriarch of the family, and everybody would come to me, as a bigger sister here.’ (African/Caribbean female; distressing anxiety, sought formal help, received medication)*

> *‘You see, there’s one lady who sits with us at the club and nothing’s ever right for her, and you realise that if you’re like that, people say, “Oh, I don’t want her sitting next to me,” do you know what I mean? Because she talks negative.’ (White British female; non-distressing anxiety, used self-help only)*

South Asian and African/Caribbean participants tended to be especially concerned about gossip.

> *‘Interpreter: …they [her friends] won’t keep it a secret and then they’ll just gossip about her.’ (South Asian female; distressing anxiety, used self-help only)*

African/Caribbean participants with salient religious identities relied heavily on their religious institutions for mental health support. Besides the power of collective prayers, participants with distressing anxiety appreciated the opportunity to be referred to a *‘trained therapist’* when needed and to join support groups established by their local Church to receive and offer reciprocal support from others with similar experiences.

> *‘In our area, we have those ‘home-help’, now if we live in the same community, you come together, you fellowship together. So in that process…I state my problem, then I am told, you know, there is somebody else like this, so when you meet all together, you come to see maybe you share the same problem or their problem is worse than yours, or me, I can encourage them a bit, how I used to go to counselling or something like that.’ (African/Caribbean female; distressing anxiety, sought informal help, received counselling)*

#### 4.2.3 Formal help (i.e., healthcare providers) for mental health concerns

Participants’ accounts suggest that the decision to seek formal help is influenced by a complex interplay of factors: the perceived need for formal help, conflict with higher-order goals, candidacy for treatment, and beliefs about treatment providers and different treatments.

Overall, participants believed that those who were *‘not able to cope with daily life’* should seek help. Inability to cope was conceptualised as experiencing significant, *‘long-term’* distress and impairment in important areas of functioning and, to a lesser extent, as exhibiting *‘erratic’* or harmful behaviours, such as risk to self or others, and alcohol or substance abuse.

Participants with distressing anxiety who sought formal help, which was often delayed, tended to use poignant words or phrases, such as *‘felt absolutely awful’, ‘desperation’*, *‘miserable’*, *‘suicidal’* and *‘crisis’,* to describe their emotional state at that point. Interestingly, participants with distressing anxiety who experienced symptoms suggestive of comorbid depression tended to seek help primarily for depression rather than anxiety. Likewise, participants who differentiated depression from anxiety believed that experiencing depression, as opposed to anxiety, was an indication to seek formal help.

The perceived need for formal help was also identified by close, trusted individuals, who played a significant role in either enabling or deterring participants with distressing anxiety from seeking treatment.

> *‘…my elder daughter, she had said, “Dad, I think it would really help you to talk to someone.” (African/Caribbean male; distressing anxiety, sought formal help, received counselling)*

However, perceiving a need for help did not always translate into formal help-seeking behaviour, as this decision entailed a dynamic process. It involved balancing the perceived need for help and beliefs about candidacy for treatment and weighing them against higher-order goals. This balance shifted throughout the participants’ illness journey. Participants believed that health services should be reserved for those with serious, objective physical signs of illness, a conceptualisation that did not encompass mental health problems. Furthermore, participants who prioritised projecting a *‘strong’* self-image and interpreted help-seeking for mental health concerns ‘*as a sign of weakness*’ delayed or refused to seek help. Among these participants, males tended to prioritise upholding the *‘typical man’* social image over seeking help, ‘*We’re supposed to be the strong ones.’* However, once achieved, this balance could lead to proactive help-seeking.

> *‘For some time I’d been working myself up to see a GP and I thought, well, you know, what’s he going to think of me? But no, he was, I thought he’d be such, sort of,*

> *‘What’s he worried about when I’ve got people here with terminal illnesses?’ But no, he was glad I came, and we had quite a long chat.’ (White British male; distressing anxiety, sought formal help, but refused treatment)*

#### 4.2.4 Beliefs about treatment providers

Across all cultural groups, most participants considered general practitioners (GPs) the primary gatekeepers to formal mental healthcare services. However, their views varied concerning the role of GPs in treating anxiety and mental health problems in general. Some believed GPs were well-equipped to manage common mental health problems, while others contended that their role should be limited to referring people to *‘trained therapists’*, psychologists or psychiatrists. Conversely, participants who had chronic physical health conditions, such as Parkinson’s Disease, expressed the importance of specialist nurses and, to a lesser extent, specialist doctors, in the early detection of mental health problems and subsequent referral to appropriate health services.

Across all cultural groups, participants’ evaluations of the quality of healthcare provided by their GP were polarised, with the majority inclining towards a negative view. Participants who did not seek help for mental health concerns often had preconceived ideas about GPs’ propensity to overprescribe antidepressants as a *‘quick fix’* for mental health concerns rather than investigate more complex underlying issues. Consequently, this discouraged them from seeking help.

Those who sought formal help explained how access to GP appointments was limited during the COVID-19 lockdown. This was further complicated by an unacceptable triaging process conducted by non-medical personnel, *‘how can a receptionist triage a patient? It’s beyond my ken.’* These barriers forced participants to seek other sources of support.

Even when offered a GP consultation, long waiting times and sudden transition to phone consultations were considered unacceptable. Time constraints during GP consultations meant that participants had to prioritise physical health problems over their mental health concerns. Furthermore, participants who were not consistently seen by the same GP often perceived GPs as dismissive. This lack of continuity of care was viewed as a barrier to appreciating the person’s narrative and taking account of their unique experiences and needs.

> *‘There’s one thing, every time you go to the GP there’s a different doctor. If it’s one that okay, they know your background, but if every time new face, different one.’ (South Asian female; distressing anxiety, sought formal help, but refused treatment)*

South Asian and African/Caribbean participants reported experiencing discrimination in healthcare settings based on their cultural background, which was frequently associated with implicit assumptions regarding their socioeconomic status.

Participants reported that racial stereotyping and lack of compassion, understanding and respect by GPs adversely affected the quality of care they received. In some cases, the substandard care and distress associated with these discriminatory experiences aggravated the participants’ mental health concerns.

> *‘…some GPs will treat someone from a minority community less respectfully than maybe someone from the White community because the assumption made is, you know, in some socioeconomic wards of [name of city] where we know literacy rates are poor, [South Asian] men or women, when they go to the GP, or it could be a Middle Eastern lady or man struggling with English will be dismissed by the GP, you know, because they don’t know the background.’ (South Asian male; non-distressing anxiety, used self-help only)*

African/Caribbean participants noted that society’s disregard for older adults was not limited to general social interactions, but extended into healthcare settings. They feared that this would negatively impact the quality of healthcare services they would receive in the future.

> *‘But I don’t want to be, when I’m old be classed, I am old, right, but to be put with elderly, to be segregated.’ (African/Caribbean female; distressing anxiety, sought formal help, received medication and counselling)*

However, participants’ views about GPs were not unanimously negative. A limited number of participants believed their GP, particularly when consistent, helped them understand their symptoms and provided a diagnosis that allowed them access to different services.

#### 4.2.5 Beliefs about treatments

##### Beliefs about medications

Across all cultural groups, participants conflated different classes of psychotropic medications, including antidepressants, benzodiazepines, antipsychotic medications and beta-blockers. Most participants believed medication should be reserved for those with *‘severe mental health issues’*, especially depression. This perspective was particularly relevant when depression was attributed to *‘more of a chemical thing than a mental thing’*.

A limited subset of participants believed that medication could be used for anxiety, but only as a *‘last resort’* if it causes prolonged significant distress, functional impairment or is complicated by other serious symptoms, such as psychosis. Participants with distressing anxiety who prioritised the projection of a strong personal identity tended to regard the use of medications for anxiety as a sign of personal weakness. Among them, those who required medication grappled with the conflict between their need for medication and how they interpreted this need.

> *‘I used to be very strong and I had strong willpower, you know, I can manage. I have so many problems in my life but I can manage really. But, when my husband passed away, after that I am getting weak and weak and weak, psychologically and emotionally and physically. Then this COVID came and suddenly, I don’t know where from, you know. So obviously I think I want to try more but I can’t. I think maybe need ((pause)) but I don’t know how to cope with it [anxiety], you know, without medicine.’ (South Asian female; distressing anxiety, sought formal help, received medication and counselling)*

Medication was generally depicted as *‘a temporary remedy’* for anxiety that mainly provides symptom relief, but *‘doesn’t solve the problem’.* Nevertheless, participants across all cultural groups had a predominantly negative view of psychotropic medications, expressing general, *‘Any drugs you take, they all have a side-effect’*, as well as specific concerns. The most feared side effects were drowsiness and dependence. Medications were believed to *‘slow you down’* and make you *‘like a bit of a zombie’.* Participants highlighted their fear of becoming ‘*dependent’* on *‘the happy tablets’,* which they perceived as *‘addictive’*. This perception was often reinforced by the notion of withdrawal symptoms following the abrupt cessation of medication. Such beliefs served as barriers to accepting medications as viable treatment options, even among those who were currently taking them. Other side effects included drug-drug interactions, especially in the context of polypharmacy, drug-food interactions, fatigue, dry mouth and gastrointestinal disturbance.

Participants evaluated the perceived necessity of medications and their potential benefits against possible adverse effects, including their impact on the self, to determine whether to initiate medication. To overcome their concerns, some participants took their prescribed medication irregularly, used lower than the recommended doses or asked their GP to avoid the *‘stronger’* medications.

Essentially, they were testing the medications, weighing their effectiveness in controlling symptoms against side effects to determine what they considered to be the optimal dose [24]. Those who passively accepted medication tended to be older adults with distressing anxiety and comorbid depression. Participants were generally uncertain about when the medications were expected to take effect and how long they should be taken.

##### Beliefs about psychological therapies

Participants tended to limit talking therapies to counselling. Those who preferred counselling over other treatments believed that *‘opening up’* to a therapist about their problems, revisiting traumatic experiences and having the opportunity *‘to talk through things’* could help alleviate their burdens. Participants expected therapists, as professionals, to guide them through their recovery journey and empower them rather than offer predetermined solutions. Therapists were *‘detached’* from family and social circles, which facilitated the process of disclosure.

> *‘You lighten yourself to strangers you don’t know.’ (South Asian female; distressing anxiety, sought formal help, received medication and counselling)*

Conversely, participants who criticised counselling argued that it fails to address the root causes of mental health concerns and instead redirects one’s focus to the past rather than the future. These participants were also sceptical of sharing private problems with *‘strangers’*, preferring to keep their problems to themselves or discuss them with trusted family members, friends or, in the case of the African/Caribbean group, their *‘pastor’*.

> *‘Probably, I feel it’s [the therapist] a stranger, I know they’re professional people, but I feel that it’s a stranger, I’m going to talk about my personal life, I’d rather talk to who I’m close to, like I said my mother, my big sister, them the people I feel they’ll understand.’ (South Asian male; distressing anxiety, sought formal help, received medication)*

The effectiveness of talking therapies, in general, was believed to depend on the individual’s motivation to engage in therapy*, ‘If the person doesn’t want to help himself, nobody can help’*, and the therapist being a knowledgeable, supportive, non-judgmental, active listener who could *‘click’* or *‘gel’* with the client. The importance of client-therapist matching on the basis of gender, culture and religion was only expressed by South Asian participants with salient cultural and religious identities.

Only White British participants with distressing anxiety recalled that they had received cognitive behavioural therapy (CBT). Nevertheless, participants from all cultural groups had an unclear understanding of CBT. Those who did not receive CBT tended to confuse it with counselling, while those who received CBT considered it a *‘tool to use’* that should be frequently administered and struggled to remember what it involved. CBT was believed to be an intervention that does not take into account the needs of different populations.

> *‘So, we have a huge push by organisations or medical professionals to promote CBT as a main line of intervention. It may work for, again I hate to use the word, it might work for a ‘middle class White professional’, but sad to say, it probably will not work as good as an intervention for people whose English language is poor, have low literacy levels or skills.’ (South Asian male; non-distressing anxiety, used self-help only)*

Group therapy was welcomed by participants who believed it could offer an opportunity to share experiences, gain practical advice and feel that they are not alone. However, it was perceived as unacceptable by those who were concerned about privacy and the negative impact listening to other people’s problems could have on their mental health.

## 5 DISCUSSION

This multicultural qualitative study explored and compared beliefs underlying coping strategies for anxiety among self-reporting White British, South Asian, African and Caribbean older adults in the UK, using Leventhal’s Common-Sense Model [10]. It incorporated the perspectives of 27 older adults with distressing anxiety and 25 with non-distressing anxiety. Across all cultural groups, participants adopted different strategies to manage anxiety, including self-help and informal and formal help-seeking. Their strategies shifted in response to their evolving illness journeys. Self-help strategies were fundamental to all participants. However, the decision to seek help was determined by a complex interplay of factors not recognised by Leventhal’s Common-Sense Model [10]. Notably, older adults’ salient identities, rather than their cultural background, influenced their selection of coping strategies.

Our findings suggest that older adults predominantly use self-help strategies to manage anxiety. Some strategies appear to offer temporary relief by targeting negative thoughts and anxiety symptoms, while others have more long-lasting effects by leveraging older adults’ self-resources, such as cherished personal attributes (i.e. personal identities), social roles (i.e., role identities) and group memberships (i.e., social identities) [25]. By capitalising on these self-resources, older adults could shift their focus from their vulnerabilities towards distinct, unaffected, positive and valued aspects of themselves. That enabled them to broaden their self-view and restore an overarching positive self-image [26].

The emphasis on self-help strategies echoes the findings of previous systematic reviews on depression [27, 28] and anxiety [4] in older adults. This is especially significant considering that older adults in our study believed that inability to cope, manifested as significant distress and functional impairment, was the only indication for formal help-seeking, but primarily associated this with depression rather than anxiety. Underrating anxiety, even among those experiencing distressing anxiety, could be attributed to older adults’ fragmented understanding of these disorders. Findings from our previous qualitative analysis [13] have highlighted a disparity in older adults’ awareness of anxiety and depression. While older adults view depression as a serious, uncontrollable illness that requires treatment, they have an incoherent understanding of anxiety - they neither normalise it nor consider it as an illness trajectory. This awareness gap could partly explain the lower rates of mental health services use among older adults with anxiety compared to those with depression [3].

Faced with these challenges, one could argue that public education campaigns are necessary to improve older adults’ awareness of anxiety disorders and available support from healthcare services. However, we would emphasise that, given the prominence of self-help strategies and the unclear understanding of anxiety disorders in this group, older adults do not only need public awareness campaigns, but also need interventions that would enable them to tap into their self-resources and deploy self-help strategies more effectively. These interventions could be viewed as more acceptable adjuncts or, in mild cases, alternatives to formal therapeutic support.

One such group of interventions is social prescribing services that connect individuals, mainly through referrals from voluntary organisations and self-referrals [29], to non-clinical, community-based services [30]. Much of its practice relies on helping people engage in activities and social groups. Our study highlights that meaningful, goal-directed activities and social groups serve a function beyond mere pleasure; they provide older adults with avenues to affirm positive and desirable aspects of their identity [31]. Moreover, older adults who strongly identified with their groups gained access to additional social and psychological resources - valuable social networks, a sense of self-esteem, belonging, meaning and purpose. This aligns with the extensive body of “social cure” research, which suggests that internalised, positive group memberships act as social cures promoting well-being and health [32]. This was particularly evident among South Asian and African/Caribbean participants who had salient religious and cultural identities and joined groups characterised by these interlinked, harmonious identities.

Older adults sought informal support from trustworthy, empathetic family members and friends. Quantitative evidence on the effect of received social support on health outcomes is decidedly mixed [33]. One explanation for this inconsistency is that received support is not highly correlated with perceptions of support [34] (i.e., perceived availability of and satisfaction with social support), a subconstruct of social support that has been consistently linked with positive health outcomes [35, 36]. The characteristics of the support provider and provider-recipient relationship identified in our study could help clarify the factors required to translate received support into perceived support.

Additionally, our findings suggest that a complex interplay of factors influences an individual’s decision to seek formal help. While Leventhal’s Common-Sense Model [7, 10] primarily focuses on illness representations, proposing that coping behaviours (e.g., formal help-seeking) directly follow from the evaluation of health threats (e.g., anxiety), it overlooks the challenges of taking action and the role of social influences on taking action [37]. Our study suggests that the perceived need for formal help-seeking, as recognised by the participants and close members in their social networks, influences older adults’ willingness to seek help. Perceived need could be understood as a personal threshold at which functional impairment and emotional distress resulting from anxiety surpassed the older adult’s subjective (or objective, as observed by others) ability to cope.

Candidacy for care, a construct originally introduced by Dixon-Woods, Cavers (38), describes how individuals negotiate their eligibility for healthcare. It appears that older adults tend to view serious, objective physical signs of illness as legitimate grounds for seeking help, unlike mental health problems that tend to lack observable signs. Additionally, beliefs about treatment providers and pharmacologic and psychologic treatments, shaped by previous experiences or anticipated concerns, impacted the participants’ help-seeking behaviour.

Notably, a basic premise of Leventhal’s Common-Sense Model [39] is that individuals are motivated to select and perform coping behaviours to reduce a perceived discrepancy between their current state and a desired goal (i.e., not experiencing anxiety). However, this narrow focus on a single goal fails to consider the complexity and multiplicity of goal structures [40]. Indeed, specific goals are embedded within complex hierarchies, where higher-level, more abstract goals related to one’s identity are pursued through lower-level, more concrete goals.

Moreover, at any one time, individuals have multiple goals that differ in importance, that may either facilitate or interfere with one another [41]. Conflicts between goals may arise due to the mutual incompatibility of the behaviours needed to attain them [40]. In our study, prioritising a strong or positive self-image sometimes precluded participants from seeking help. It was not the participants’ cultural background that dictated this prioritisation, but rather their higher-order identity goals.

Older adults have multiple salient identities, including personal, role and social identities. These identities are dynamic and contextually defined; they interact and intersect in complex ways. Overall, older adults tended to adopt identity-congruent coping behaviours. Their salient identities, rather than their cultural background, influenced their selection of coping strategies. This complexity rendered older adults across all cultural groups more similar than different. The conception of culture as a relatively static, invisible force that divides people into distinct, internally homogeneous groups and shapes their decisions and beliefs portrays individuals as passive recipients devoid of individual agency [42]. It neglects within-group differences by disregarding the crucial role of one’s multifaceted self-identity in interpreting self-relevant events, selectively adopting cultural patterns and regulating behaviour. Cross-cultural research should embrace the diversity and heterogeneity of individuals both within and across groups rather than homogenise entire cultural groups.

### Strengths and Limitations

This study is the first multicultural qualitative inquiry to explore older adults’ beliefs about coping strategies for anxiety. A significant limitation is the recruitment of a self-reporting sample and the use of varied terms to describe anxiety. Our public contributors expressed concern that relying solely on Western medical terminology, such as “anxiety”, might imply that only individuals who had been clinically diagnosed or those who had previously sought help from healthcare services were eligible, thereby excluding potentially eligible older adults with undiagnosed anxiety. This group is particularly important, as little research has explored their perceptions of anxiety and the beliefs underpinning their coping strategies.

Public contributors who self-identified as South Asian or Caribbean also noted that medical labels may not be widely recognisable in their communities, may not translate easily into other languages, and may carry stigmatising connotations. Therefore, the public contributors and research team agreed to include lay terms such as “worrier” and “stress” in the study flyers. While this enhanced inclusivity, it also meant that participants with a wide range of experiences could have been included, potentially introducing variability in the data. Despite implementing several measures to ensure all participants had experienced anxiety either in the past or at the time of the study, we believe that the sample included two groups of participants: those who likely met the diagnostic criteria for anxiety disorders (i.e., distressing anxiety) and those who did not (i.e., non-distressing anxiety). To reflect these differences, we developed a typology distinguishing between the two groups.

## 6 CONCLUSIONS

Interventions that empower older adults to capitalise on their self-resources and deploy self-help strategies more effectively can serve as acceptable adjuncts to formal therapy. Nevertheless, it is essential to address barriers that preclude older adults from seeking formal help, particularly among those with a perceived need to seek help. No one model can depict the complexity of coping behaviours. While applying Leventhal’s Common-Sense Model yielded novel insights, it could not fully capture the motivational factors underlying one’s decision to seek help. Cross-cultural research should acknowledge heterogeneity within groups rather than impose boundaries of proposed homogeneous entities if it is to provide nuanced and accurate insights.

## Data Availability

The anonymised dataset is available for restricted data-sharing to authenticated researchers who provide verifiable institutional affiliation and have ethical approval in place. A metadata record of the anonymised dataset is available on Figshare (https://doi.org/10.48420/28468466) for further information and guidance.

## 7 ACKNOWLEDGEMENTS

We are extremely grateful to the participants for giving us the unique opportunity to learn from their experiences. This research would not have been possible without their generosity and willingness to share their experiences. We extend our heartfelt thanks to our public contributors, whose insights and collaboration have significantly enriched this project. We are also indebted to the community and charity organisations that agreed to host our arts-based workshop and presentations, and those that helped us access a broader audience of potential participants: Older Black Africans Day Opportunities (OBADO), Khush Amdid, Yellowbird Age Friendly, Hulme Carer’s Forum, African Caribbean Care Group, Parkinson’s UK, Stroke Association Carers Group, Growing Old Disgracefully, GM Older People’s Network and Dementia United.

This work was supported by the President’s Doctoral Scholar Award (a PhD studentship awarded by The University of Manchester, United Kingdom).

